# The FLuorometholone as Adjunctive MEdical Therapy for Trachomatous Trichiasis Surgery (FLAME) Trial: Study Design

**DOI:** 10.1101/2024.06.26.24308549

**Authors:** Ahlam Awad Mohammed, Aida Abashawl, Sarity Dodson, Wondu Alemayehu, Alemu Gemechu, Aemero Abateneh Mengesha, Dereje Kumsa, Tony Succar, Yineng Chen, Kathleen McWilliams, Vatinee Y. Bunya, Maureen G. Maguire, Matthew J. Burton, Gui-shuang Ying, John H. Kempen, FLuorometholone as Adjunctive MEdical Therapy for Trachomatous Trichiasis Surgery (FLAME) Trial Research Group

## Abstract

**Purpose:** To report the design of <u>FL</u>uorometholone as <u>A</u>djunctive <u>ME</u>dical Therapy for TT Surgery (FLAME) Trial.

**Design:** Parallel design, double-masked, placebo-controlled clinical trial with 1:1 randomization to fluorometholone 0.1% eyedrops twice daily or placebo twice daily for four weeks in eyes undergoing trachomatous trichiasis (TT) surgery; assessing the efficacy, safety, and cost-effectiveness of fluorometholone 0.1% in preventing recurrent postoperative trichiasis.

**Methods:** Up to 2500 eligible persons with trachomatous trichiasis (TT) undergoing lid rotation surgery will be enrolled in Jimma zone, Ethiopia. Participants, surgeons, study field staff, and study supervisors leading operational aspects of the trial are masked to treatment assignment. Randomization is stratified by surgeon, which simultaneously stratifies by the district. The study visits are at baseline/enrollment, at four-week post-enrollment, six months, and one year (study exit). The primary outcome is cumulative one-year postoperative TT (PTT) incidence, defined as: ≥1 lashes touching the globe, evidence of epilation, and/or repeat TT surgery. Secondary postoperative outcomes include number of trichiatic lashes, location thereof (touching the cornea or not), evidence of post-operative epilation, entropion, changes in corneal opacity, IOP elevation, need for cataract surgery, visual acuity change from baseline, eyelid contour abnormality, granuloma, eyelid closure defect, and occurrence of adverse events. Health economic analyses center on calculating the incremental cost per case of PTT avoided by fluorometholone treatment.

**Conclusion:** The FLAME Trial is designed to provide evidence of the efficacy, safety, and cost-effectiveness of adjunctive topical peri-/postoperative fluorometholone 0.1% therapy with trichiasis surgery, which is hypothesized to reduce the risk of recurrent trichiasis while being acceptably safe.

Trial Registration: ClinicalTrials.gov # NCT04149210

## INTRODUCTION

Trachoma is the leading infectious cause of blindness worldwide.^1-2^ Trachomatous trichiasis (TT), a sequela of conjunctival scarring resulting from repeated infection/chronic inflammation, is a key mechanism leading to blindness, and also causes severe, chronic eye pain. Approximately 1.9 million people are visual impaired from trachoma, including 0.45 million who are irreversibly blind; as of April 2023, it was estimated that approximately 1.5 million people have untreated TT and are at risk of blindness in addition to having eye pain.^3^ TT surgeries are performed by integrated eye care workers (trained nurses) as part of the permanent public health system to speed up the TT backlog clearance.^4^

Surgery to relieve trichiasis is one of the four World Health Organization (WHO)-endorsed “SAFE” Strategy priority interventions for programs aiming to prevent trachoma blindness.^1^ The elements of the strategy, under the acronym “SAFE,” include: <u>S</u>urgery for trichiasis, <u>A</u>ntibiotics to clear *C. trachomatis* infection, <u>F</u>acial cleanliness to reduce *C. trachomatis* transmission, and <u>E</u>nvironmental improvement to remove conditions facilitating endemic trachoma. Unfortunately, a high recurrence rate following trichiasis surgery limits the benefits of surgery and also reduces community confidence in the surgery, resulting in less utilization by people who could benefit from it. Reports of the incidence of recurrence of TT following TT surgery (“post-operative TT” [PTT]) have been highly variable, and are hard to combine due to heterogeneous methods used in reporting. In a comprehensive review summarizing reports through 2012, the median for the currently WHO-recommended procedures (bilamellar tarsal rotation and posterior lamellar tarsal rotation) appears to be in the 20-30% range, with programmatic results tending to be less favorable than clinical trial results which often involve merit-based selection and intensive training of TT surgeons.^5^ Reports based on selection of the best TT surgeons followed by intensive training in a clinical study, such as a trial, would not be expected to be representative of typical programmatic conditions.

Reported risk factors for PTT include surgeon skill, severity of preoperative disease, older patient age, and ongoing inflammation during the perioperative period.^5-12^ Optimization of surgical quality is acknowledged as critical for obtaining good results of surgery, and a subject of considerable programmatic effort, but such efforts have not eliminated recurrence.^13^ PTT is a difficult problem, with worse outcomes of surgical repair than primary TT.^14^ According to WHO, repeat TT surgery should be performed by an ophthalmologist, but ophthalmologists rarely are available in the impoverished communities typically afflicted by trachoma. Because of the dire situation of patients who have PTT—with high risk of blindness, ongoing pain, and limited management options— and the impact on community observations of such cases in reducing interest in sight-saving TT surgery, avoidance of PTT is of paramount importance.

Ongoing inflammation in the setting of trachoma is associated with progressive conjunctival scarring,^15^ and often is seen in persons with TT.^8,16-17^ Such inflammation is only rarely associated with active *C. trachomatis* replication,^18^ and the specific causes thereof are incompletely understood. In the STAR Trial, where azithromycin therapy was associated with reduced risk of postoperative TT even though detectable *C. trachomatis* infection was rare prior to treatment, anti-inflammatory effects of azithromycin were cited as one potential mechanism by which benefit may have occurred.^7^ Considering the potential importance of ongoing perioperative inflammation in relation to PTT incidence and the rarity of active *C. trachomatis* replication among TT cases, members of our group previously undertook a randomized phase 2-like clinical trial evaluating whether perioperative use of a low potency, low cost corticosteroid eyedrop (fluorometholone 0.1%) might improve outcomes. In evaluating three potential doses, we found all three were associated with an approximate one-third reduction in PTT compared with placebo-treated eyes and with contralateral untreated eyes; the safety profile appeared favorable.^19^

Because the phase 2-like trial was not sufficiently powered to definitively prove topical fluorometholone 0.1% is effective in reducing the incidence of PTT, we have undertaken a full-scale clinical trial adequately powered to settle the issue, described here: the FLuorometholone as Adjunctive MEdical Therapy for TT Surgery (FLAME) Trial. The specific aims of the FLAME Trial are: 1) To assess the efficacy of fluorometholone 0.1% one drop twice daily for four weeks in reducing the incidence of post-operative TT when given as adjunctive therapy with TT surgery in the programmatic setting; 2) To assess whether such treatment is sufficiently safe for wide-scale implementation in TT programs; and 3) To estimate the costs of adding fluorometholone 0.1% treatment to TT surgery per case of postoperative TT averted, and to characterize the value of such treatment under a range of plausible health economic circumstances.

## METHODS

### Overview of trial design

The FLAME study is a National Eye Institute-funded randomized, double-masked, parallel-group, placebo controlled clinical trial (Clinicaltrials.gov # NCT04149210) in which a total of up to 2,500 eligible study participants with TT undergoing lid rotation surgery (table 1) are enrolled to assess the efficacy and safety of fluorometholone 0.1% as ancillary therapy for trachomatous trichiasis (TT) surgery. The Trial’s location is in the Jimma zone of the Oromia Region of Ethiopia, wherein the SAFE strategy for trachoma blindness mitigation is being implemented (including the conduct of TT lid rotation surgery for a large volume of patients). Study participants are randomized in a 1:1 ratio to fluorometholone 0.1% versus placebo (artificial tears). Participants may have one or both eyes enrolled in the trial; when both eyes are enrolled the participant uses the same treatment for both eyes. Patients are examined at baseline, four weeks post-surgery, six months, and one year at trial exit (see Table 2). Before recruiting any study participants into the study, written approval of the protocol and of the informed consent form were obtained from all governing Institutional Review Boards (IRBs), and approval maintained thereafter. The study participants’ confidentiality is protected by providing each eligible participant an enrollment ID, and locking away all documents containing identifiers, such participants’ consent forms, contact information forms, and enrollment logs. The Ethiopian IRBs include the National Research Ethics Review Committee of the Ministry of Education (formerly under the Ministry of Science and Technology), and the Ethiopia Food and Drug Administration. The Oromia Health Bureau also reviewed and supported the study. In addition, IRB approval was obtained from the Study Chair’s institution (Massachusetts Eye and Ear under Mass General Brigham, Boston, Massachusetts, USA) and the Vice-Chair’s institution (London School of Hygiene and Tropical Medicine, London, United Kingdom). As per National Institutes of Health “single IRB” requirements, the Data Center Director’s institution (University of Pennsylvania) formally ceded responsibility for IRB approval to Mass General Brigham’s IRB.

### Study organization

The study group consists of four resource centers: the Chairman’s Office, the Data Center, the Field Coordinating Center, and the Surgical Team. The Chairman’s Office provides overall leadership to the FLAME Trial Research Group mentioned in the credit roster. The Field Coordinating Center (FCC) oversees the operational aspects of the study in a manner typical for a clinical trial coordinating center, except for handling the data system, data management, and statistical aspects of the study which is managed by the Data Center. The FCC oversees field implementation of the trial in Ethiopia, employing, training, and supervising all field team members, except those involved in TT programmatic services under the Surgical Team. The Surgical Team oversees the implementation of TT surgery, including conducting TT patients’ mobilization (through mass mobilization and house-to-house visit), and presenting TT patients to the FCC study field team for consideration of enrollment into the study. The Data Center oversees and manages the data system and conducts data management, and statistical analysis for the study.

### Study treatment

The study treatments are fluorometholone 0.1% or placebo, each one drop two times daily for four weeks. The study test articles are prepared by the Penn Investigational Drug Service, under the Data Center in Philadelphia, repackaging active treatment or placebo into identical bottles and shipping them to Ethiopia. Fluorometholone 0.1% suspension was donated by Allergan, now operated by Abbvie (North Chicago, Illinois, USA) (FML^®^ Ophthalmic Suspension, USP 0.1%). Artificial tears (Soothe XP^®^ artificial tears produced by Bausch and Lomb) were purchased; both have a similar milky appearance. Shelf-life of the repackaged drug was assessed by testing a sample of bottles at six, twelve, and eighteen months after preparation, and it was determined to be potent (90-110% of expected potency) for at least twelve months, after which study drug is considered expired. The packaging contains a medication box number corresponding to the masked contents of the bottle, on a peel-off label. Each box number maps to the masked treatment group; all are unique so as to prevent unmasking of a large segment of the study population should unmasking of one participant’s treatment be required.

Study participants are asked to complete a diary provided by study field team regarding eyedrop use and to bring the diary and their medication bottle at the week four visit. Adherence is assessed by questioning patients at the week four visit, by tabulating the number of eyedrop administrations recorded in the diaries, and by evaluating the difference in weight of the eye drop bottles from baseline to four weeks using scales (OHAUS^TM^ Navigator^TM^ Portable Balance, Model: NV222) able to detect a difference in weight of 0.05 g (the approximate weight of one eyedrop).

### Study participant selection and eligibility criteria

Patients who have TT confirmed by the study’s Surgical Team, who plan to undergo upper lid rotation surgery conducted by a participating surgeon, and who potentially are interested in participating in the study, formally provide informed consent to undergo screening procedures to determine eligibility (Table 1). Those who are eligible are invited to participate in the study and go through the enrollment process, get examined, and be randomized into the Trial. Both participants and those who elect not to participate undergo TT surgery as per standard of care.

### Surgical treatment

Since the study treatment with fluorometholone 0.1% would be added to existing programs if cost-effective, the goal of the study is to assess the impact of adding fluorometholone treatment to an unaltered existing programmatic system. Therefore, the study does not seek to change the practices of the participating surgeons or the standard operating procedures of the extant TT surgery outreach program. In this region, new surgeons are taught to perform Posterior Lamellar Tarsal Rotation (PLTR or “Trabut”), which was found superior to Bilamellar Tarsal Rotation (BLTR) surgery in a randomized trial reported during the conduct of our preliminary trial,^18^ after which the WHO endorsed PLTR as an approved procedure for TT surgery as part of the SAFE program. However, the WHO still recommends BLTR for surgeons who are more familiar with BLTR. Participating surgeons use whichever of these two procedures they prefer. Thirty surgeons, previously trained and assigned to work in various district of the Jimma zone but with varying number of years of experience ranging from less than a year to over six years, have participated in the FLAME Trial. Among these surgeons, 23 perform PLTR surgeries and seven surgeons use the BLTR surgical method. Dissolvable sutures are used exclusively.

### Visit procedure and schedule

Study participants are scheduled to be seen four or five times in total. After a baseline visit at the time of enrollment (Visit 1) and a surgical visit (Visit 2), which typically occur on the same day, the study participant is scheduled to return for follow-up at four weeks (day 26-35) (Visit 3), 6 months (+/- 60 days) (Visit 4), and one year (+/- 90 days) (Visit 5) after enrollment/trichiasis surgery (Table 2). After completing the one-year visit, study participants exit the study. The study participants’ contact information and residence location is collected at baseline, and each participant is given a patient ID card with visit dates to facilitate follow-up visit procedures.

Per the extant TT program, separate from the trial, surgeons are expected to examine TT patients after their surgery at various times such as the first postoperative day, after 8-14 days, at three months. Limited data about complications are collected at those clinical care visits.

### Randomization procedures

A surgeon-stratified randomization schedule was prepared in advance by the study Data Center. Randomized patients either immediately undergo the Visit 2 procedures (which includes trichiasis surgery), or else can be scheduled to return to the center for Visit 2 within seven days.

Package boxes of 20 medication kits are prepared by the Investigational Pharmacy in using random block sizes of two or four in such a way that every package box is comprised of 10 fluorometholone and 10 placebo bottles. A specific package box is used by each surgeon, who works through one box at a time, thus achieving stratified randomization by surgeon. Surgeons operate in their own geographical area. Thus, this method of stratification also ensured there would be an approximately balanced number of patients in each treatment group within each geographical area. When package boxes are incompletely used by a given surgeon, they can be repackaged per instructions from the unmasked biostatistician at the Data Center to create a new package box with 10 active and 10 placebo treatment bottles so as to avoid wasting treatment bottles.

At the surgical location, after obtaining consent and confirming eligibility, the study team enters the patient’s name and date of enrollment on the Enrollment Log for study patients of each participating surgeon. A pre-selected enrollment ID is assigned to each sequential patient. A medication box number is assigned to each enrolled patient following a randomization sequence, and the medication box kit number label is put on the enrollment form, thus accomplishing randomization at enrollment. Therefore, each patient has a screening ID number (one of each of these is given to each patient screened, including those not ultimately enrolled, for purposes of constructing a CONSORT Diagram). In addition, each enrolled patient receives a patient ID number and uniquely identified medication box kit. Study participants are counted as enrolled in the study, and part of safety and efficacy analyses, from the moment the patient’s name is entered in the Enrollment Log. Medication box number(s) are used to link to the simultaneously issued actual treatment assignment for the statistical analysis of efficacy and safety data.

After treatment assignment, the study Field Team proceeds to implement the treatment protocol for Day 0, treating the study eye(s) with an eye drop from the assigned treatment kit prior to surgery. The treatment kit contains a bottle with sufficient fluorometholone 0.1% or placebo for all study eyes to receive four weeks’ treatment. The study participants are carefully instructed on how to apply eye drops, how often to apply them, and for how many weeks. Participants practice using an eyedrop from eyedrops from single-use preservative-free artificial tear containers and the drop from the assigned medication bottle. After receiving their first assigned eye drop treatment just prior to surgery, study participants undergo TT surgery, and then resume their study treatment upon removal of any bandages and eye patches the following day at the Day 1 postoperative follow-up performed by the TT surgeons.

### Outcome Assessments

Assessments of outcomes and other study variables are measured primarily during eye examinations. Efficacy assessments included trichiasis and entropion grading. Postoperative TT is defined as at least one of the following: (1) one or more lashes touching the globe in an eye; (2) evidence of epilation on clinical examination; and/or (3) history of repeat TT surgery. Additional assessments include clinical signs of trachoma status, visual acuity, patient-reported outcomes and elements of cost calculations as mentioned in the section regarding health economic analysis. Adverse outcome assessments include examining corneal scarring, assessing and documenting the presence of lagophthalmos, and other conditions such as overcorrection, lid margin notching, granuloma, blepharitis, necrosis of eyelid margin, IOP elevation, occurrence of cataract surgery (within the study period), infectious keratitis, and other observed adverse events attributed to study treatment.

Eye examinations are conducted by masked, trained and study-certified field team nurses, using +2.50 diopter magnifying loupes and a torch. The outcome assessment and examination details along with the visits are given in Supplementary Appendix 1.

The primary outcome of the FLAME Trial is the cumulative incidence of PTT as determined by the trained study team members at four weeks, six months and one year. In addition, safety outcomes, patient-reported outcomes and health economic issues are evaluated. Safety outcome measures include intraocular pressure (IOP) elevation, cataract, adverse events attributed to study treatment. Patient-reported outcomes include overall patient satisfaction, cosmetic outcome, impact on quality of life, Ocular Surface Disease Index^15^, and the five-level EQ5D and thermometer EuroQol Questionnaire^16^. The latter primarily uses a new official translation into the Oromo Language, although pre-existing Amharic and English versions also are available.

### Quality Assurance

This study is monitored by various study team members at the FCC, Chair’s Office and Data Center using different procedures to assure protocol adherence and high quality data. The specific quality assurance features related to data integrity of the FLAME Trial are:

- Standard data collection forms and procedures;
- Standard electronic case reporting data system (REDCap) with the built in functions for checking of data ranges, consistency and completeness;
- Masking of study participants, surgeons, and all study staff in Ethiopia to the assigned treatment group of the study participant;
- Training and certification of study coordinators in data collection and data entry;
- Explicit instructions with the release of new/revised data collection forms about new/revised questions and instructions;
- Frequent checking of data mainly by the Data Center biostatistician, but also by the field teams, supervisors, internal monitors to detect any data problems early and provide feedback to the field teams for any correction;
- Timely data detection (e.g., range checks) and edits for missing, invalid, and unexpected (potentially erroneous) responses;
- Regular reporting on performance (timeliness, completeness and accuracy of data) of field teams for the data collection;
- Regular field observation and review of data collection process by study officers and team supervisor, including reexamining certain study participants
- Checking by the study supervisors, field study teams, and internal monitors of a random sample of entered data by the field teams against original source data after data editing has been completed.
- Supervision of the Trial by a Data and Safety Monitoring Committee (DSMC) which is an independent board appointed by the National Eye Institute (NEI) as advisory to the FLAME Trial Research Group and the NEI. The DSMC meets approximately biannually and is responsible for ongoing review of the efficacy and safety data, policy and ethical issues, and study performance.

### Sample Size

Prior to the enrollment, the sample size was estimated for comparing the fluorometholone 0.1% treatment group and placebo control group for the cumulative TT recurrence rate by 12 months at type 1 error rate of 0.05. Assuming TT recurrence by 12 months of 20% in the placebo group, and 15% in the fluorometholone group (a 25% reduction in TT rate), 75% participants with bilateral TT surgery, inter-eye correlation coefficient of 0.48 in TT recurrence^18,20^ and 10% lost to follow-up, a total sample size of 2254 participants (3944 study eyes) provided 90% power for comparing difference using repeated measures logistic regression for per-eye analysis with the inter-eye correlation adjusted by generalized estimating equation (GEE).^21^ However, during enrollment the bilaterality proportion was found to be considerably lower than 75%, so sample size re-calculation was performed using the observed bilaterality rate of 35% as well as a slightly lower observed one-year cumulative incidence of postoperative TT in the placebo group (slightly less than 19%), and the observed lower rate of loss to follow-up (2%), which provided a total sample size target of 2400 participants for enrollment. A revised sample size of approximately 2400 was approved by the DSMC.

The number of eligible participants for enrollment and the number of eligible eyes required are in the following Table 3 for the assumed PTT rate ranging from 17% to 20% in the placebo group. Because in a field trial simultaneously enrolling large numbers of participants daily at sites often off the cellular grid, it is not possible to reach an exact number and stop, a target enrollment of up to 2,500 participants (1,250 participants per group) was selected; the revised total approved by the study IRBs.

### Statistical Analysis Plan

Since some participants undergo concurrent TT surgery in both eyes, and their treatment outcome (i.e., recurrence of TT postoperatively) is correlated, the primary assessment of efficacy is the comparison of cumulative proportion of incident post-operative TT by 12 months between the two treatment groups using a repeated measures logistic regression model, where the inter-eye correlation is accounted for through generalized estimating equations. The difference for cumulative incidence rate of postoperative TT by 12 months, the odds ratio and their 95% confidence intervals for the comparison between two treatment groups are calculated from the repeated measures logistic regression model. Analyses are performed with their treatment group assignment based on randomization regardless of their treatment compliance (“intent-to-treat”).

To check the consistency of results over subgroups, effect modification of the treatment on the primary outcome is assessed with the following factors by including treatment group indicator, subgroup indicator and their interaction term in the repeated measures logistic regression model as described above. If we find any important interactions, stratum-specific recurrence rate by treatment group and their odds ratios (ORs) for treatment effect will be reported for each effect modifying variable, which are considered for interactions.

a. Baseline upper eyelid trichiasis severity with two severity categories (severe: 6 or more in total number of upper eyelid lashes or epilation >⅓; not severe: 5 or less in total number of upper eyelid lashes or epilation <1/3).
b. Baseline conjunctival (papillary) inflammation (presence or absence)

A secondary analysis also will be performed based on the level of adherence to randomized treatment with fluorometholone or placebo. For this analysis, good treatment adherence will be defined as one of the following criteria meet:

1. Bottle weight change indicates > 75% of expected doses and the medication diary indicates > 75% of expected doses;
2. Medication diary indicates >75% of expected doses and the bottle weight change is unknown;
3. Bottle weight change indicates >75% of expected doses and the medication diary is unknown;
4. Self-reported adherence is very good, when both medication diary and the bottle weight change are unknown.

Because only small percent (<5%) of participants may be lost to follow-up or will not comply with the trial protocol during the study, the primary statistical analysis for primary outcome and secondary outcomes will be based on participants who complete the 1-year follow-up (completed cases) with their treatment group assignment classified as assigned at randomization (“intent-to-treat”). Sensitivity analyses including “per protocol” analysis and missing data imputation will be performed to assess the robustness of the results with respect to loss to follow-up and non-compliance with the eligibility criteria and the treatment protocol.

### Health Economic Analysis

The cost of the intervention will be calculated as the cost of the drops of fluorometholone 0.1% that are dispensed to patients. The price of the generic drug will be based on what importers charge for fluorometholone 0.1% suspension in Ethiopia. The cost of post-index surgery medications related to the eye will be derived from the surgeon-reported medications for each patient. The costs to the program in Ethiopia also will be taken into account. The cost of any additional medical care utilization (not including study visits) will include utilization based on self-report and a price based on standard use by a clinic in Ethiopia. Costs of reoperations for TT specifically will be considered. All costs will be converted to US dollar values at the time of the study based on the international exchange rate. The outcomes will be cases of post-operative TT averted. If the proposed intervention is more effective and more expensive, the incremental cost-effectiveness ratio will be calculated as the difference in costs divided by the difference in postoperative TT cases. If one intervention is more effective and less expensive then it will be described as dominating and is the obvious economic choice. Quality-adjusted life years (QALYs) also will be calculated based on the change from baseline for the intervention and control group and take the difference. The incremental cost-effectiveness ratio will be expressed as dollars spent per QALY gained. Finally, sensitivity analyses varying one variable at a time will be conducted to determine whether any reasonable variation in costs or any observed variable being at the high or low end of a confidence interval rather than at the mean would change the interpretation of the economic value of the intervention (i.e., it would go from being cost-effective to not or vice versa.) If a change in one variable at a time or a change when making all variables either the most or least favorable to fluorometholone would change the qualitative interpretation of the results, the results will be bootstrapped to characterize the level of certainty about the conclusion using a cost-effectiveness acceptability curve.

## CONCLUSION

The FLAME Trial is a randomized, double-masked, placebo-controlled parallel sequential randomized field clinical trial to evaluate the efficacy, safety and cost-effectiveness of fluorometholone 0.1% eyedrops as an ancillary therapy for trachomatous trichiasis surgery in Ethiopia. The primary outcome is the cumulative incidence of postoperative TT. Additional outcomes include ocular surface outcomes, potential adverse outcomes of surgery or study treatment, patient-reported outcomes, health economic analysis, and presenting visual acuity. Preliminary data and theoretical suggestions have led us to hypothesize that fluorometholone 0.1% will be efficacious and sufficiently safe for programmatic use as well as cost-effective.^18^ The FLAME Trial is expected to conclusively confirm or refute these hypotheses. If the hypotheses are supported, adding fluorometholone 0.1% eyedrops to TT surgery programs would be justified, and might avert large numbers of cases of post-operative TT with high risk of vision loss, chronic eye pain and limited options. Based on the current backlog, reduction in postoperative TT from 20% to 15% corresponds to about 75,000 fewer people with postoperative TT globally; a larger effect size would avoid more postoperative TT cases. Additional incident cases are expected for many years into the future; lower risk of PTT also would benefit those persons with new TT cases.

## Supplemental Material

Appendix 1 – FLAME Trial Data Summary

Appendix 2 – De-identified ocular photos of study participant with trichiasis at various visits

## Data Availability

All data produced in the present study are available upon reasonable request to the authors

## ACKNOWLEDGEMENTS

The authors acknowledge National Eye Institute Program Officers Sangeeta Bhargava, PhD, and Jimmy Le, ScD.

## Appendix 1 FLAME Trial Data Summary

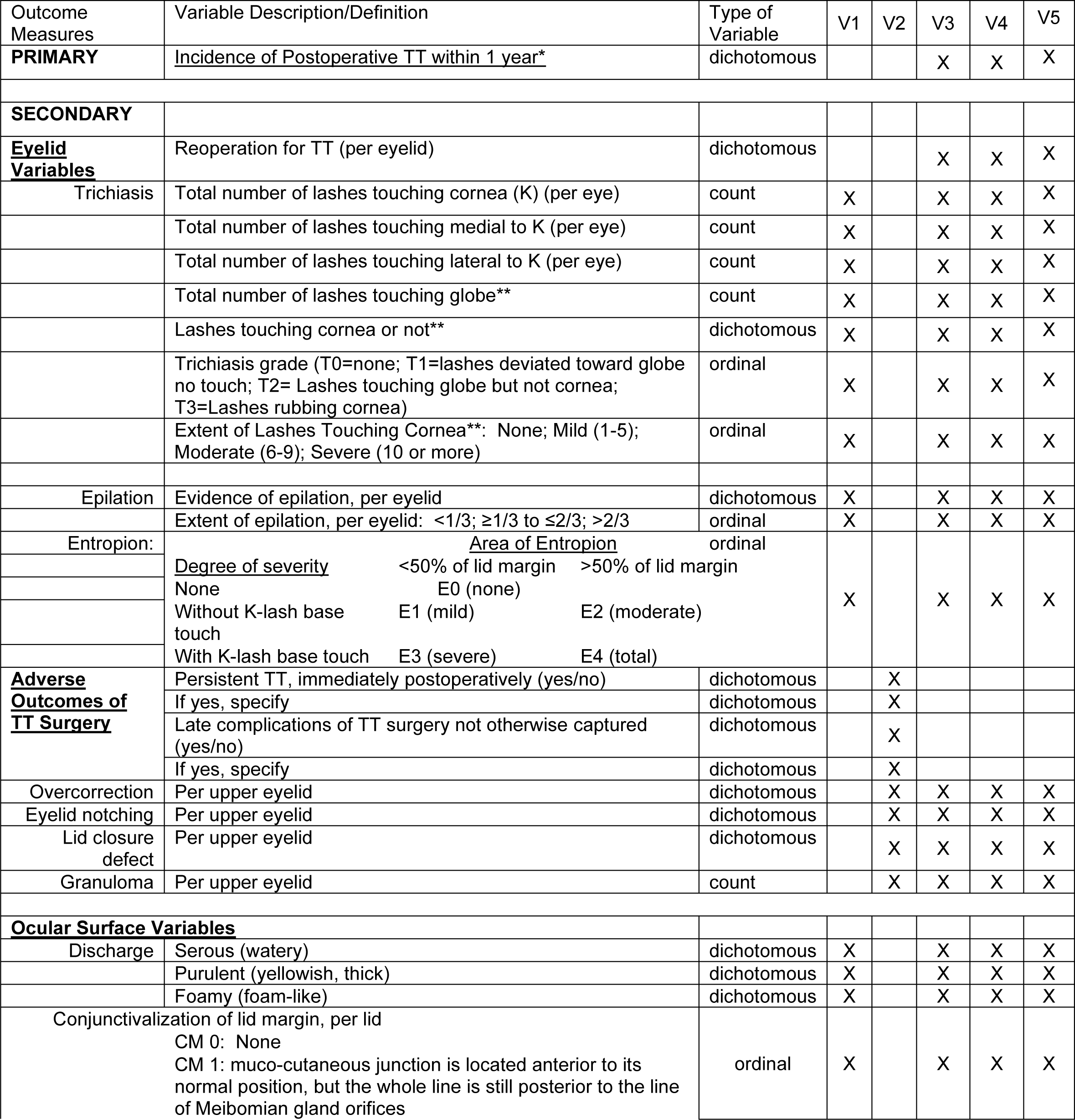
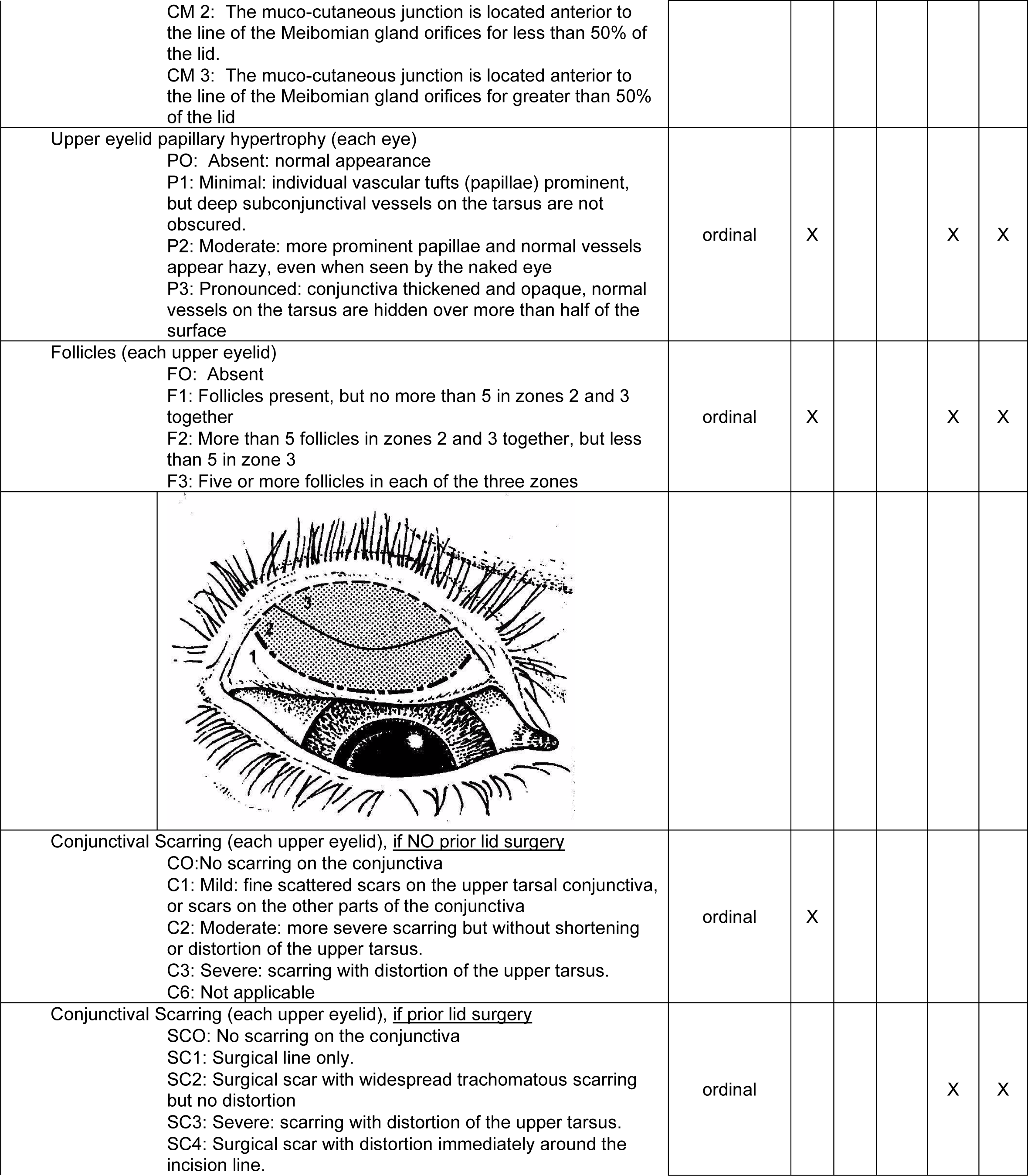
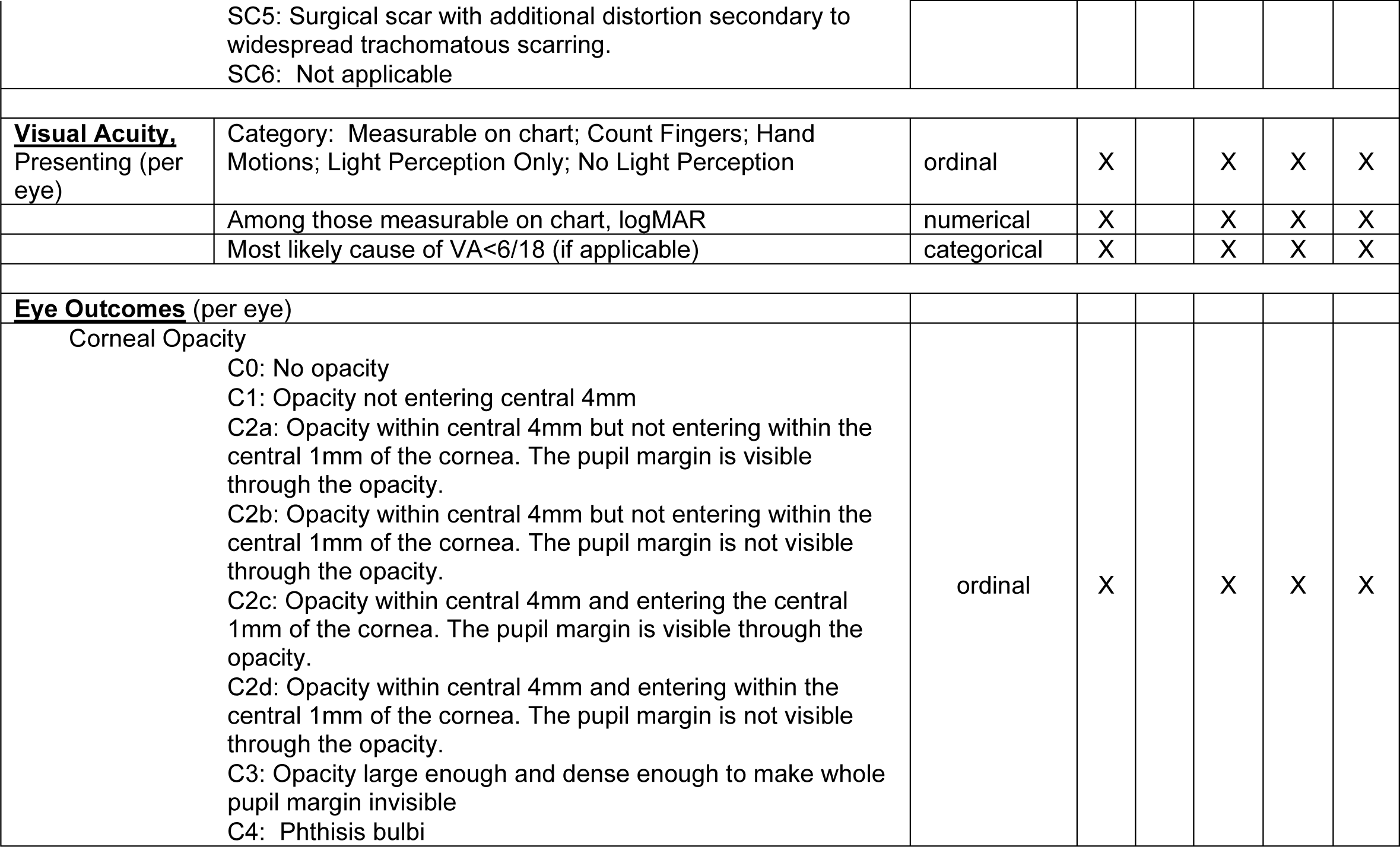

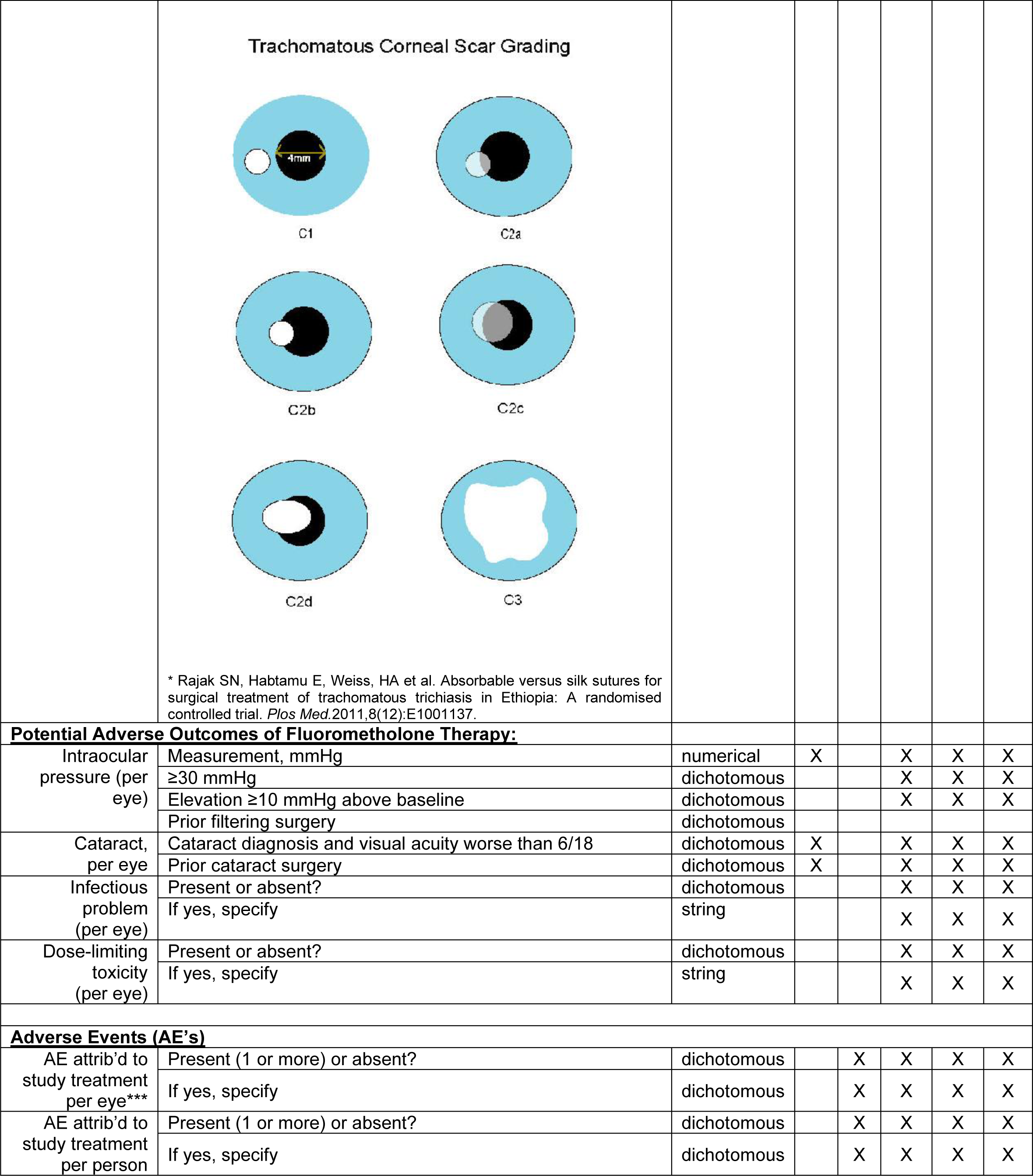
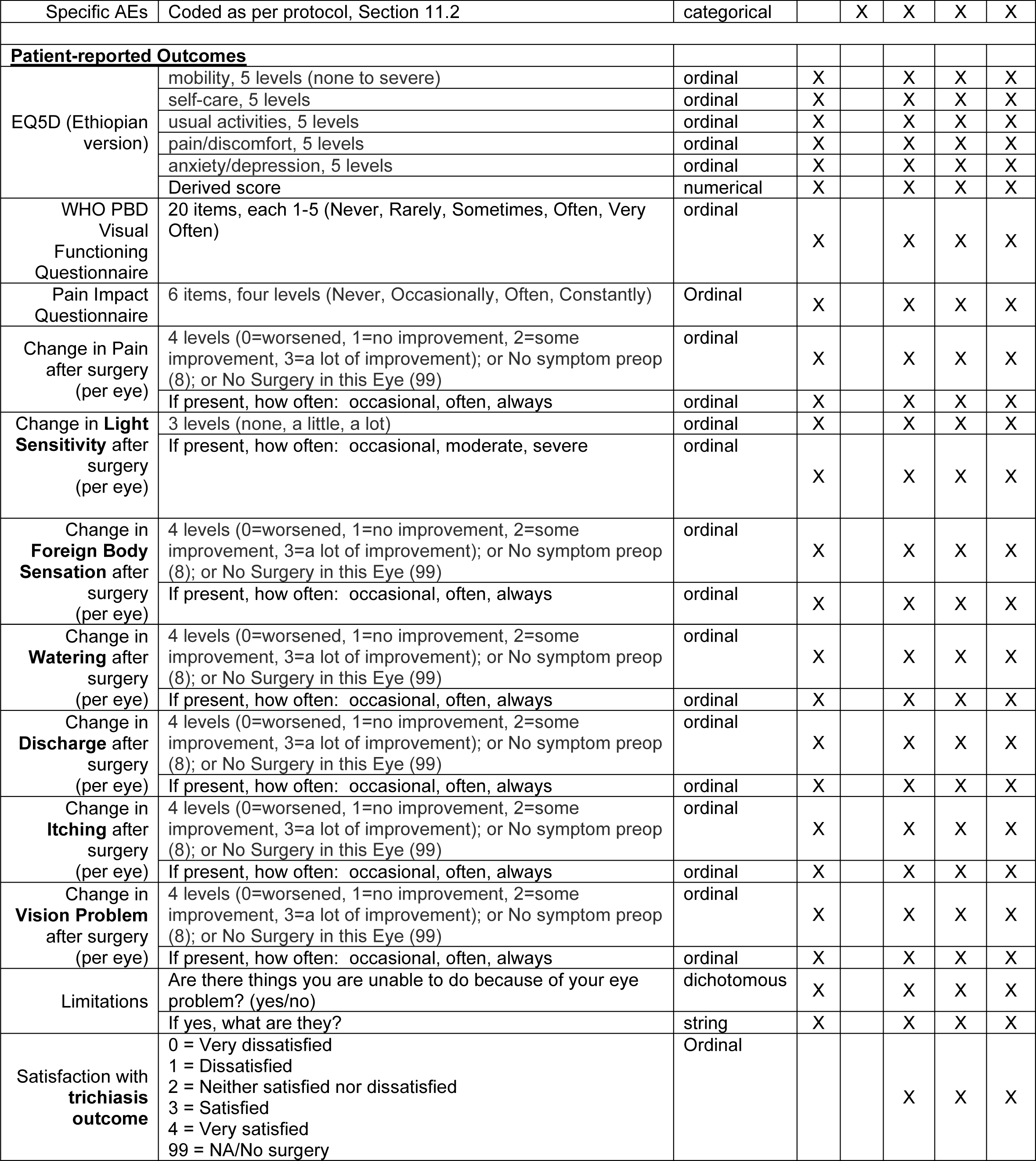
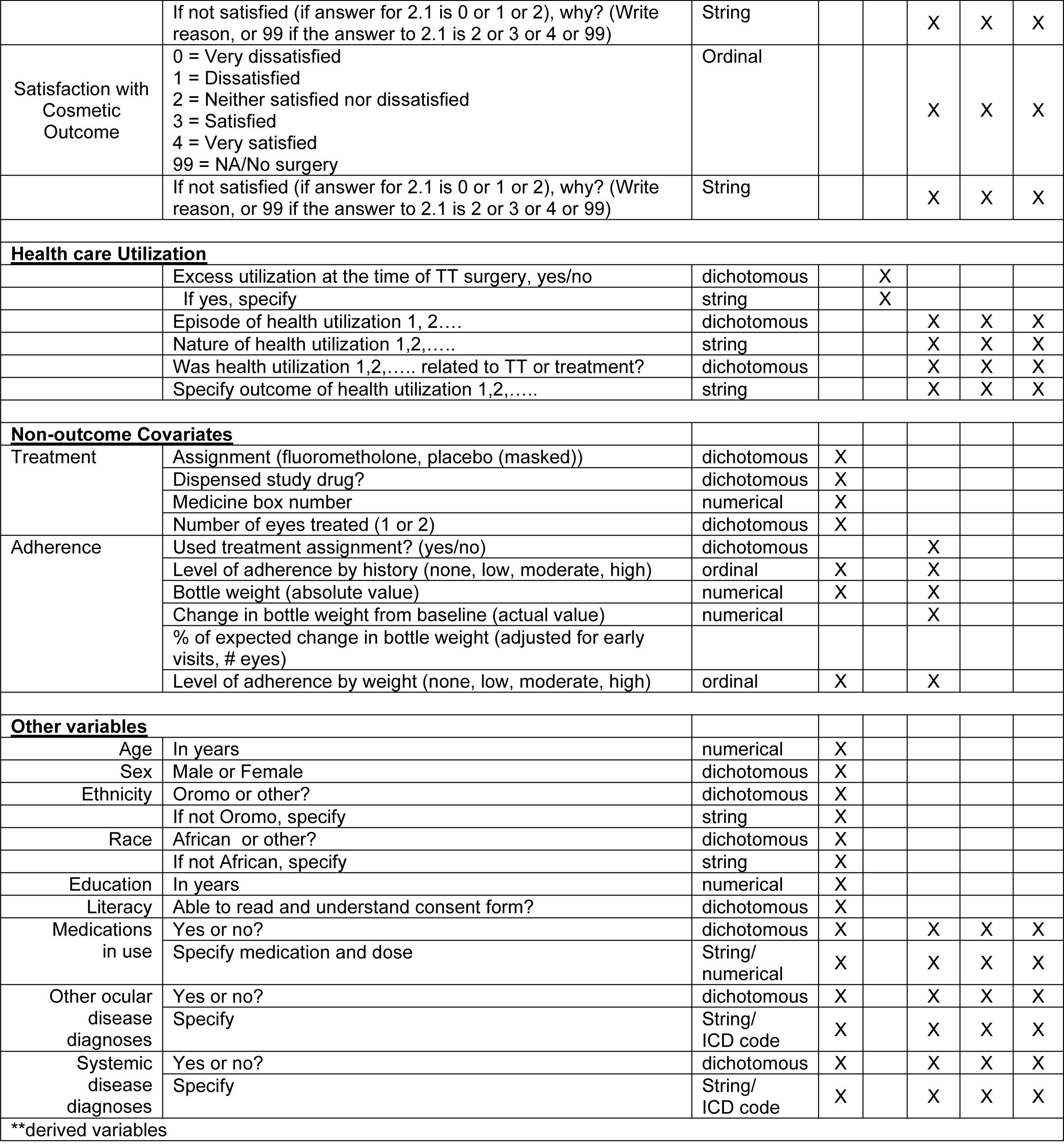

